# Black bone MRI morphometry for mandibular cortical bone measurement in head and neck cancer patients: Prospective method comparison with CT

**DOI:** 10.1101/2020.07.21.20154880

**Authors:** Joint Head and Neck MRI-Radiotherapy Development Cooperative, Lisanne V. van Dijk, Juan Ventura, Kareem Wahid, Lin L. Zhu, Brigid A. McDonald, Sara Ahmed, Keith Sanders, Sonja M. Stieb, Lance McCoy, Christina S. Sharafi, Kathryn Preston, Joly Fahim, Aubryane DeArmas, Mona Arbab, Yao Ding, Jihong Wang, Bastien Rigaud, Anando Sen, Mark Chambers, Katherine A. Hutcheson, Kristy K. Brock, Stephen Y. Lai, Abdallah S. R. Mohamed, Clifton D. Fuller

**Affiliations:** Department of Radiation Oncology, The University of Texas MD Anderson Cancer Center, Houston, TX, USA; University Medical Center Groningen, University of Groningen, Groningen, NL; The University of Texas MD Anderson Cancer Center UTHealth Graduate School of Biomedical Sciences, Houston, TX, USA; University of Houston, College of Medicine, Houston, TX, USA; Prairie View A&M University, Prairie View, TX, USA; Department of Radiation Oncology, Indiana University, Indianapolis, IN, USA; Department of Radiation Physics, The University of Texas MD Anderson Cancer Center, Houston, TX, USA; Department of Imaging Physics, The University of Texas MD Anderson Cancer Center, Houston, TX, USA; Department of Head and Neck Surgery, The University of Texas MD Anderson Cancer Center, Houston, TX, USA; Department of Molecular and Cellular Oncology, The University of Texas MD Anderson Cancer Center, Houston, USA; Program for Image-guided Cancer Therapy, The University of Texas MD Anderson Cancer Center, Houston, TX, USA

**Keywords:** Osteoradionecrosis, Bisphosphonate-Induced Osteonecrosis of the Jaw, Magnetic Resonance Imaging, Cortical Bone, radiation

## Abstract

**Objectives:** To determine the utility of low-flip angle “black bone” magnetic resonance imaging (MRI) for cortical mandibular bone assessment by comparing interdentium cortical measurements and inter-observer morphometric variability in relation to computed tomography (CT).

**Methods:** Quantification of cortical mandible bone width was performed as per Hamada et al. at 15 cross-sectional interdentium locations on pre-treatment black bone MRI and CT for 15 oropharyngeal cancer patients, with inter-observer analyses on a subset of 3 patients by 11 observers. Bland-Altman limits of agreement and bias estimation, Lin’s concordance correlation (LCC), and Deming orthogonal regression were used to compare CT and MRI measurements. The absolute variance and intraclass correlation coefficient (ICC) were implemented for the inter-observer error quantification.

**Results:** Both the Bland Altman and Deming regression analyses showed CT and black bone MRI measurements were comparable within ±0.85 mm limits of agreement, and systematically smaller for MRI. LCC (0.60[0.52;0.67]) showed moderate equivalence between modalities. The average absolute variance between the observers was similar on CT (1.13±0.06 mm) and MRI (1.15 ±0.06 mm). The ICC analyses showed that measurement consistency was significantly higher (p<0.001) for the black bone MRI (0.43[0.32;0.56]) than CT (0.22[0.13;0.35]); nonetheless, the ICC was poor for both modalities.

**Conclusions:** Black bone MR sequence is usable as an alternative to CT for cortical mandible bone measurements, allowing use for early detection of cortical alteration (e.g. osteonecrosis). The cortical bone measurements showed substantive but equivalent inter-observer variation on both CT and black bone MRI. (Semi)automated measurement may mitigate this in future work.

## Introduction

Osteonecrosis of the mandible is a severe morbidity that has violent impact on the quality of life of patients following administration of either specific medication or radiation [1–7]. Medication-related osteonecrosis of the jaw (MRONJ) occurs in 0.1-15% of patients following bisphosphonates (BPs) and denosumab (Dmab) administration [8,9]—likely the result of poor mucosal wound healing due to the effect of medications on osteogenic cells, osteoclasts, human fibroblasts, and angiogenesis [1,3]. Similarly, osteoradionecrosis (ORN) occurs in 1-11% of the head and neck cancer patients following radiotherapy [7,10]. As such, early diagnosis and intervention, whether conservative or surgical, are crucial to improve outcomes for patients at risk of developing MRONJ or ORN. However, at present, there exists limited imaging criteria for subclinical cortical bone alteration as a correlate/surrogate of osteonecrosis.

High-frequency imaging is required to comprehensively map the development of osteonecrosis over time and to identify patient-specific imaging biomarkers. This has particular value in ORN, as regions of mandible adjacent to tumor volumes are comparatively more exposed to radiation. The demand for frequent imaging contradicts the desire to minimize the exposure of patients to additional non-uniform radiation dose. As part of a programmatic effort to evaluate longitudinal observational image biomarkers of radiation-associated mandibular injury, we are conducting large scale multi-time point MR-based imaging in order to understand the natural history and pathophysiology of ORN. Herewith, we have sought to incorporate MRI-based morphometry to characterize interval changes in cortical bone as a potential surrogate for CT imaging, which carries a non-trivial radiation dose with each additional CT scan[11].

Quantitative measurement of cortical mandible bone alteration on CT images, as proposed by Hamada et al., is performed by measuring the cross-sectional width of the cortical bone at 15 interdentium mandibular spaces as well as the density of the cancellous bone [12]. They showed that this method could significantly differentiate between patients who developed MRONJ and those who did not, as well as an additional control group. Iwata et al. also showed significant differences between MRONJ cases using CT metrics, albeit subject to relatively large inter-observer variability [3].

A promising MRI sequence, the low-flip angle or “black bone” acquisition developed by Eley et al., may be a good candidate to serve as an alternative to craniofacial imaging with ionizing methods (i.e. CT) [13,14]. Black bone MRI sequences aim to suppress fat and water simultaneously to create uniform signal for soft tissues, thus differentiating it from bone tissue with high-spatial resolution for applications in the head and neck such as craniosynostosis, skull trauma, or 3D-printing [15–18]. However, while Eley *et al*. showed that black bone MRI has comparable spatial performance for CT measurement across the image, the feasibility and utility of using black bone MR images for quantitative cortical assessment for the mandible has yet to be explored.

The purpose of this study was to perform a formal methodological comparison of black bone MRI cortical mandible bone morphometry with matched CT at the same time-point for head and neck cancer patients before start of treatment, as well as to simultaneously perform a comprehensive quantification of inter-observer morphometric variability for both imaging modalities.

## Methods

### Patients and scanning parameters

Pre-treatment black bone MRI and CT scans were acquired from 15 oropharyngeal cancer patients in a head and neck thermoplastic mask as part of a larger prospective ORN trial (NCT04265430) aiming to investigate ORN development on serial MR imaging. There was no tumor involvement into the mandible for patients included in this study, and all patients had an Eastern Cooperative Oncology Group performance status of 0-2. All patients gave written informed consent for CT.

Nine patients received the black bone sequence on a 3T Discovery MR 750 (GE Healthcare, Waukesha, MI) from July 2015 to July 2016, and 6 patients underwent a similar sequence on a 1.5T MAGNETOM Aera scanner (Siemens Healthcare, Erlangen, Germany) from May 2017 to January 2018. Repetition and echo time were slightly higher for the Siemens acquisitions (Table 1). Intensities were scaled differently, and the range of maximum MRI intensity was 1213 – 5951 for 3T GE and 169 – 203 for 1.5T Siemens scans. The CT scans were acquired with several different scanners and scan parameters (Appendix A1), with median in-plane resolution of 0.98mm (range: 0.49-1.04mm) and median slice thickness of 2.5mm (range: 1.0-3.8mm). All scans were 120kV. One patient received intravenous contrast agent.

**Table 1.** MRI scanning parameters

|  | GE<br>Discovery | Siemens<br>Aera |
| --- | --- | --- |
| Number of patients | 9 | 6 |
| Patient Weight (kg) | 69.0 – 108.0 | 60.8 – 125.6 |
| Field strength (Tesla) | 3 | 1.5 |
| TR (sec) | 6.0 – 7.1 | 8.6 |
| TE (sec) | 2.1 | 4.2 |
| Flip Angle (°) | 5.0 | 5.0 |
| Pixel Spacing (mm) | 0.5 | 0.3 |
| Slice Thickness (mm) | 2.4 – 4.0 | 1.0 |
| Slice Spacing (mm) | 1.2 – 2.0 | 1.0 |
| Field Of View (cm) | 14.4 – 40.0 | 10.0 – 40.0 |
| Imaging Frequency (Hz) | 127.7 | 63.7 |
| Maximum scan intensity | 1213 – 5951 | 169 – 203 |

### Cortical bone measurements

Measurements were performed according to method proposed by Hamada et al.[12] on the CT and black bone MR images for the 15 patients with RadiAnt DICOM Viewer software (Poznan, Poland, version 2020.1). Figure 1 details the measurement procedure. The buccal and lingual cortical bone widths were determined on a plane orthogonal to the mandible cross-section. The average of the two widths represents the final measure per interdentium location. A total of 15 interdentium spaces are measured from the left 2^nd^ molar to right. Analyses were performed 1) regarding all interdentium measurement separately, and 2) per teeth type: incisors/canines (i.e. position 0, R1-3, L1-3), pre-molars (R4-5, L4-5), and molars (R6-7, L6-7). A comprehensive practical description of the measurements is provided in the inter-observer measurement manual in Appendix A2.

**Figure 1.**
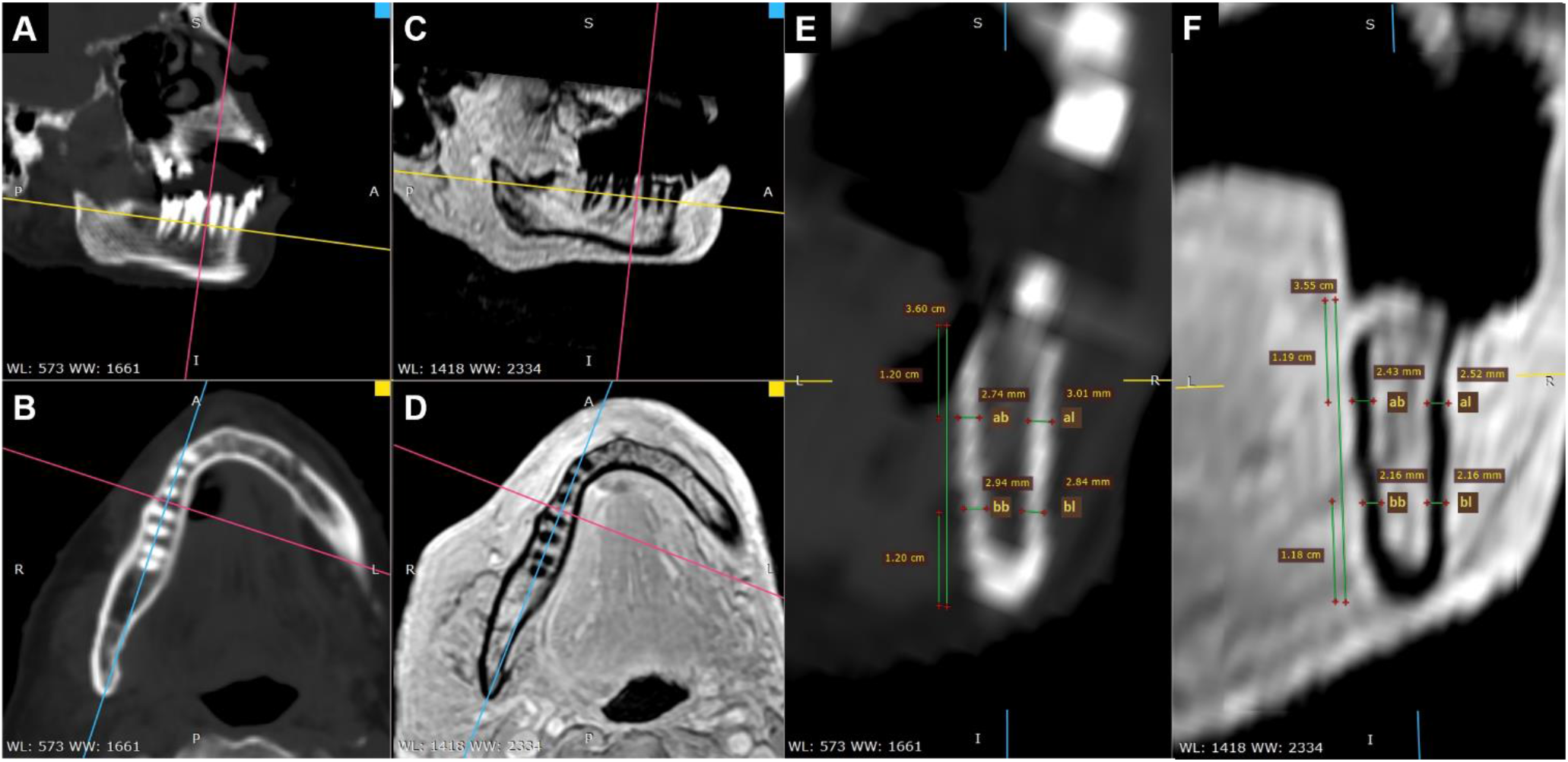
Illustration of Hamada measurement at the right interdentium space between the 2^nd^ pre-molar and 1^st^ molar for CT (A-B,E) and black bone MRI (C-D,F). The measurement is performed in the cross-sectional coronal plane (E,F) such that the sagittal (A,C) and transverse plane (B,D) are parallel to the mandible. Following Hamada et al., the height of the cortical bone measurement is defined at 1/3 and 2/3 of the length of the body of the mandible (e.g. E; total: 3.60 cm; 1/3: 1.20cm). Measuring the width of the cortical bone results in four measurements: alveolar buccal (ab), alveolar lingual (al), body buccal (bb), and body lingual (bl). Final measurement is the sum of the four individual measurements divided by 2: (ab + al + bb + bl)/2. This is repeated for all 15 interdentium spaces.

### Inter-observer analyses

Inter-observer variability analyses were conducted on a subset of 3 patients by 11 observers, which including a radiologist, a radiation oncologist and resident, a technical physician, three medical and four (under)graduate students. A manual was supplied to the observers to guide them through the CT and MRI measurement (Appendix A2). Generally, observers performed the measurements per patient on their CT and black bone MRI. Half of observers performed the measurements in reversed patient order. Observers were instructed to document measurements as well as save the measurements to JPG images (Figure 1).

### Statistical analyses

Bland-Altman limits of agreement and bias estimation, Lin’s concordance correlation, and Deming orthogonal regression were used to compare the CT and MRI measurements. Bland-Altman analysis can be used to compare two methods in the absence of a gold standard or where there is no ground truth [19]. The average between the two methods is plotted against the difference between them; the Bland-Altman limits of agreement (i.e. average difference ±1.96 times the standard deviation) is an estimate of the range within which 95% of measurement differences can be expected. The random error estimation is the standard deviation of the differences, and the systematic error estimate is the average measurement difference. The 95% confidence intervals of the biases and limit of agreement were calculated per Bland and Altman et al. [19].

Lin’s concordance analyses evaluate the equivalence of two techniques with a combination of Pearson correlation coefficient (*r*) and a bias correction factor (C_b_) [20]. Deming orthogonal regression fits a linear line between two variables (X and Y) similar to linear regression, yet differs by considering the error from both variables instead of one [21,22]. Deming regression is more appropriate when comparing two methods, as the variance of both methods is taken into account. The 95% confidence interval of the Deming regression fit was calculated with ordinary, non-parametric bootstrap resampling. Statistical analyses were performed with R packages DescTools (v0.99.35), mcr (v1.2.1), and irr (v0.84.1).

The absolute variance and intraclass correlation coefficient (ICC) were calculated for the inter-observer analyses. Variance was determined for the measurements of all observers per interdental locations, subsequently defining the mean and standard error. The ICC was performed using a two-way random effects with single rater/measurement analyses for both consistency and absolute agreement, following the Koo and Li guidelines [23]. The ICC quantifies how well observers defined the measurements to the same extent with respect to each other (agreement) or if they are correlated (consistency) [24]. In addition, the aforementioned Bland-Altman and Deming regression analyses were also performed on the measurement of the observers together.

## Results

A total of 223 CT and 210 Black Bone MRI interdentium measurements were collected from the 15 included patients. Six patients had CT metal artifacts that did not interfere with the measurements. MRI artifacts inhibited obtaining 9 measurements in one patient and 4 in another. Two MRI scans had some motion-induced geometric distortion. A single patient had a normal variant of missing a pre-molar space bilaterally, resulting in only 13 measurements for both CT and MRI for this patient.

The mean and standard deviation of the interdentium measurements over all patients was 5.89 ±0.48mm for CT and 5.61 ±0.60mm for black bone MRI. Bland-Altman analyses (Figure 2B) showed a negative systematic bias of -0.29 mm [-0.26;-0.32] for the MRI measurements compared to CT. The 95% limit of agreement was below 1 mm and the random bias was below 0.5 mm (Table 2). The Deming regression showed a significant negative intercept shift (−2.43; 95%Confidence Interval: [-3.75;-1.39]), indicating that MRI measurements were systematically lower than CT measurements (Figure 2A). The regression coefficient was 1.36 [1.19;1.58], suggesting that more deviation is seen for smaller measurements (Figure 2A). Lin’s concordance coefficient was 0.60 [0.52;0.67], indicating moderate equivalence between CT and MRI measurements.

**Table 2.**
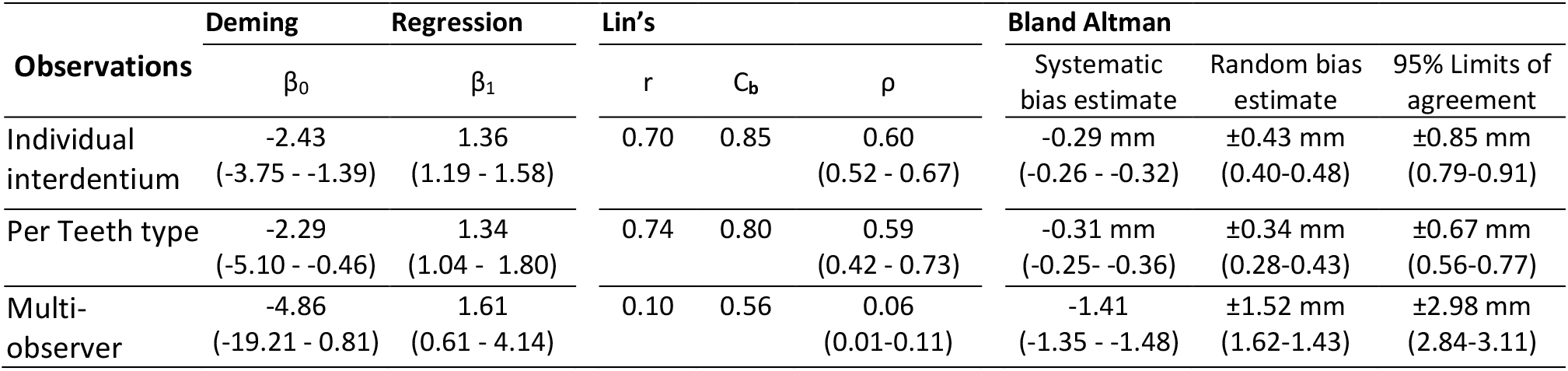
Regression and Lin’s concordance coefficients, limit of agreement and systematic random biases

| Observations | Deming | Regression | Lin's |  |  | Bland Altman |  |  |
| --- | --- | --- | --- | --- | --- | --- | --- | --- |
| | $\beta_0$ | $\beta_1$ | r | $C_b$ | $\rho$ | Systematic bias estimate | Random bias estimate | 95% Limits of agreement |
| Individual interdental | -2.43<br>(-3.75 - -1.39) | 1.36<br>(1.19 - 1.58) | 0.70 | 0.85 | 0.60<br>(0.52 - 0.67) | -0.29 mm<br>(-0.26 - -0.32) | $\pm 0.43$ mm<br>(0.40-0.48) | $\pm 0.85$ mm<br>(0.79-0.91) |
| Per Teeth type | -2.29<br>(-5.10 - -0.46) | 1.34<br>(1.04 - 1.80) | 0.74 | 0.80 | 0.59<br>(0.42 - 0.73) | -0.31 mm<br>(-0.25 - -0.36) | $\pm 0.34$ mm<br>(0.28-0.43) | $\pm 0.67$ mm<br>(0.56-0.77) |
| Multi-observer | -4.86<br>(-19.21 - 0.81) | 1.61<br>(0.61 - 4.14) | 0.10 | 0.56 | 0.06<br>(0.01-0.11) | -1.41<br>(-1.35 - -1.48) | $\pm 1.52$ mm<br>(1.62-1.43) | $\pm 2.98$ mm<br>(2.84-3.11) |

**Figure 2.**
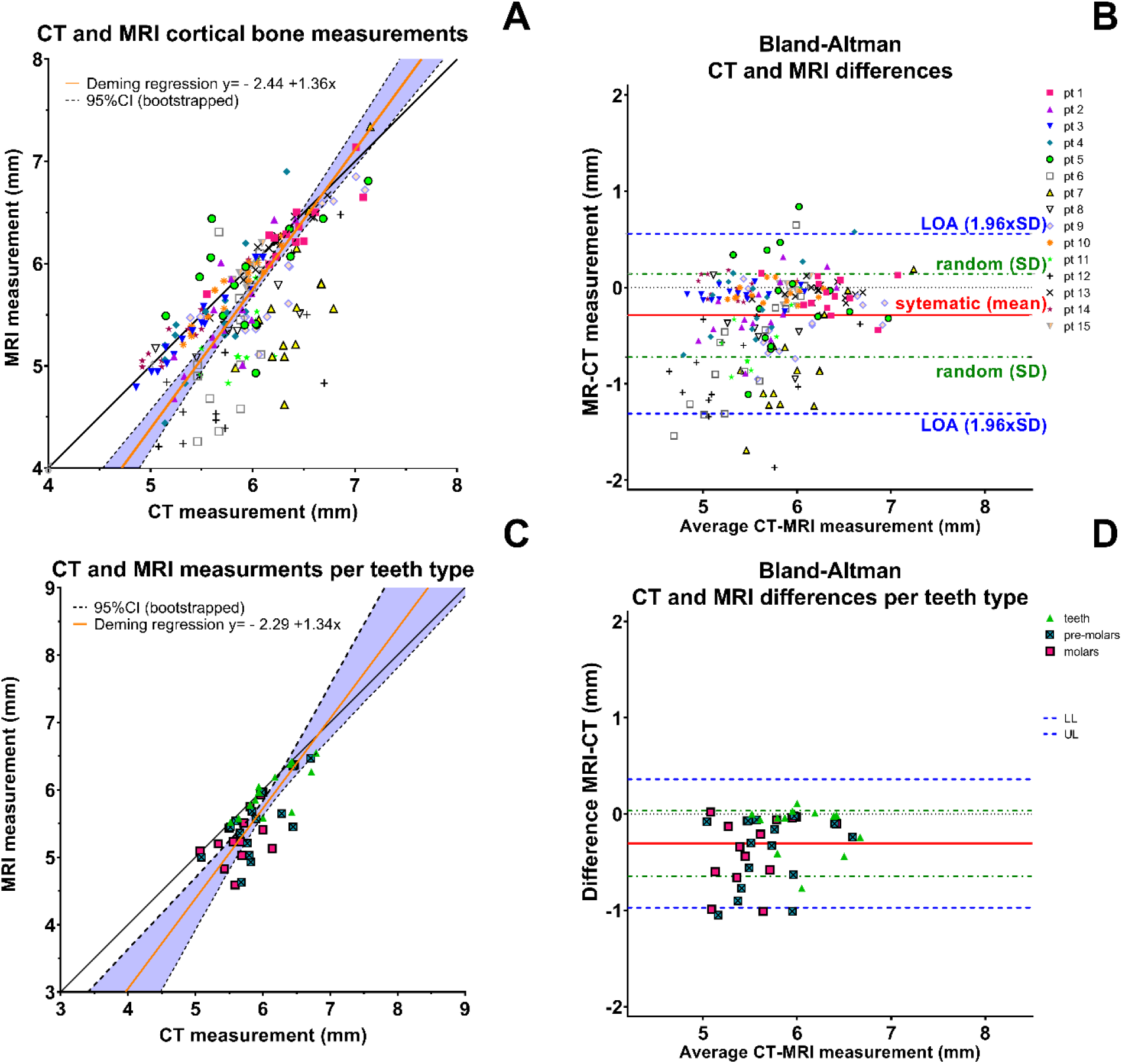
Relation and differences between cortical mandible bone width measurements on CT and black bone MRI. Scatterplot of CT measurement plotted against those on MRI and Deming regression analyses (A and C)

Averaging the measurements for each teeth type (i.e. 3 measurements per scan per patient: incisors, pre-molars, molars) showed similar regression and Lin’s concordance coefficients as the individual measure analyses (Figure 2C and Table 2). Teeth type analyses seemed to mitigate the variance between the image modalities; while fewer data points were considered, the limit of agreement and random bias were smaller (Table 2).

### Inter-observer analyses

Inter-observer data were obtained from 11 participants with 15 interdentium measurements on the CT and black bone MRI scans for 3 patients. Since 2 participants indicated that they could not complete 3 measurements, the total of number of interdentium measurements was not 990, but 984. Approximate time per scan/patient to complete the measurements was 2-2.5 hours.

The mean absolute variance and standard error of measurements between the observers on the CT (1.13±0.06 mm) were similar to those on the MRI (1.15 ±0.06 mm) (Table 3). Individual inter-observer variance per interdentium location is depicted in Figure 3. The scatterplot in Figure 4 demonstrates a sphere-like shape, indicating similar variance in CT and MRI measurements. However, the intra-class correlation for both absolute agreement and consistency was larger for the inter-observer MRI measurement than for those on CT (Table 3). This difference was significant for consistency (p<0.001), but not for agreement (p = 0.07). Nevertheless, the ICC value is poor for both modalities in agreement or consistency (ICC≤0.43), as per ICC thresholds defined by Koo and Li et al. [23].

**Table 3.** Inter-observer analyses. Mean variance between observers and intraclass correlation (ICC) analyses for agreement and consistency.

| Observations |  |  |  |
| --- | --- | --- | --- |
| Modality | mean variance (mm) | ICC agreement | ICC consistency |
| CT | $1.13 \pm 0.06$ | 0.12 (0.05-0.22) | 0.22 (0.13-0.35) |
| MRI | $1.15 \pm 0.06$ | 0.20 (0.1-0.34) | 0.43 (0.32-0.56) |

**Figure 3.**
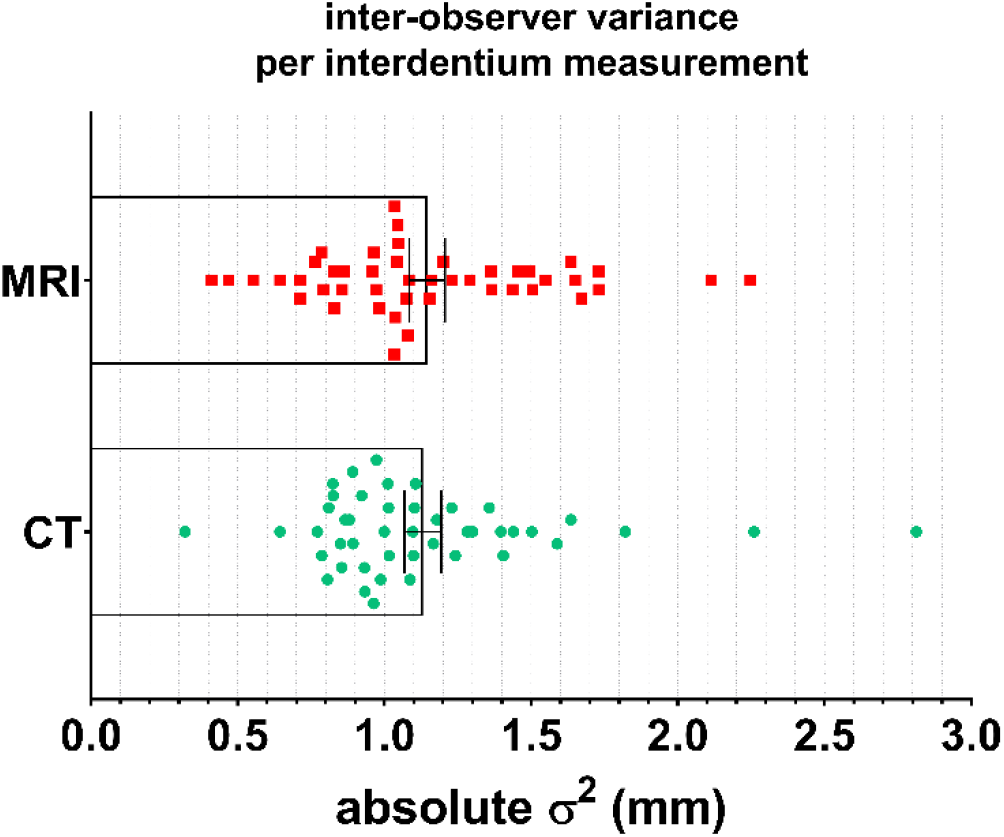
Variance of the observer measurements for black bone MRI (top/red) and CT (bottom/green). Individual points represent variance per interdentium locations (i.e. 45 total per image modality). The bar plot represents the mean and standard error (SE) of the mean.

**Figure 4.**
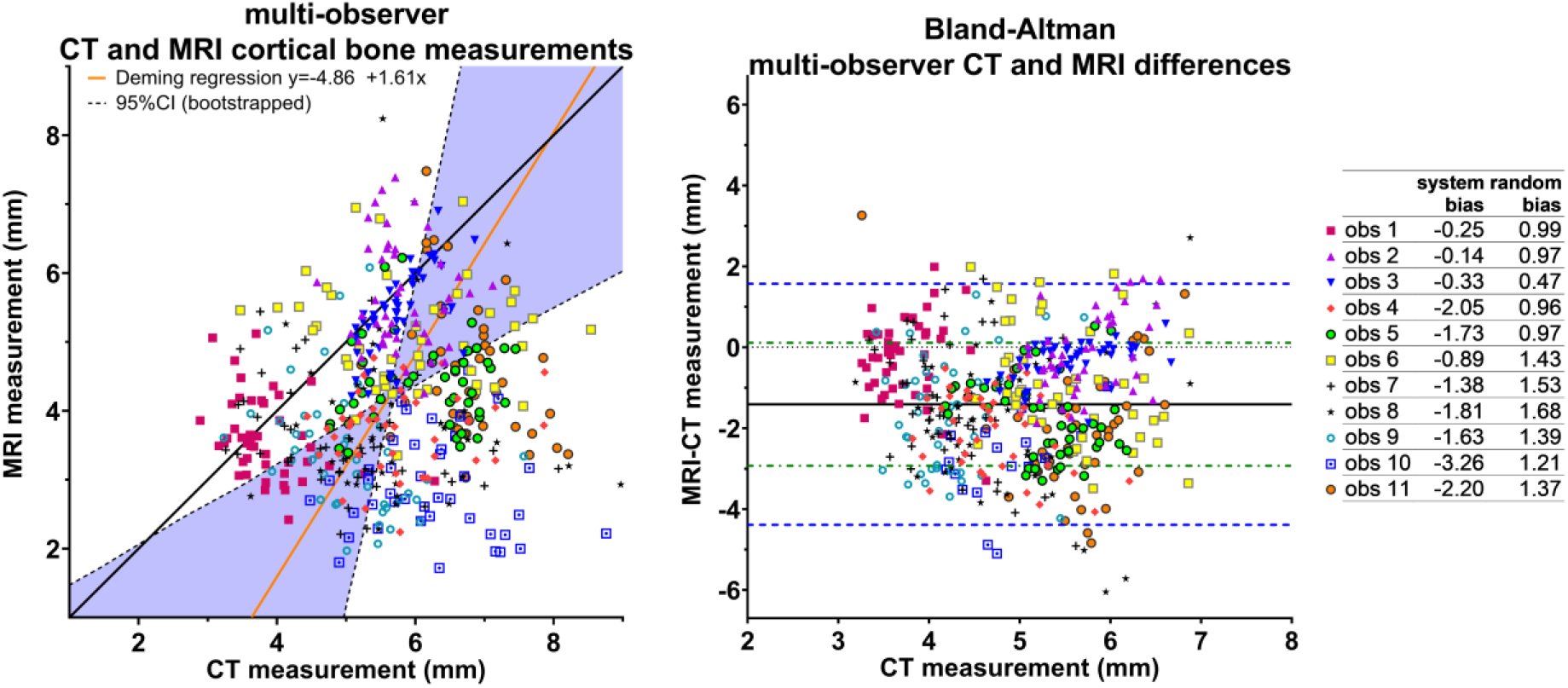
Deming regression (left) and Bland-Altman (right) plot of all interdentium measurements from the 11 observers (depicted in different colors and symbols).

The multi-observer Bland-Altman analyses also show a negative overall systematic bias of (−1.41[-1.35;-1.48]) (Table 2) as well as for all observers independently (Figure 4), indicating smaller measurements of cortical bone on MRI than on CT. Compared to the full-cohort single-observer analyses, the biases and limit of agreement between MRI and CT were increased by a factor of more than 3 times in the multi-observer analyses (Table 2), suggesting greater inter-rather than intra-observer variance. The individual bias assessment indicates that larger systematic bias component predominates with moderate random biases, suggesting again that inter-observer variability is a larger source of measurement uncertainty than modality (i.e. black bone MRI vs. CT). The Lin’s concordance also drastically decreased to 0.06 (0.01-0.11) for multi-observer measurements.

## Discussion

This is, to our knowledge, the first study to compare cortical mandible bone thickness measurements between low flip angle (i.e. black bone) MRI and the conventional CT using the Hamada method [12]. This study is a novel demonstration that MRI can be reliably used as an alternative for cortical thickness assessment in lieu of CT. This was demonstrated in 15 HNC patients, and we also subsequently determined the inter-observer variability with 11 observers in both modalities.

Our full cohort analyses indicated that—after considering a systematic correction—the CT and black bone MRI measurements are comparable within sub-millimeter limits of agreement, which is similar to the in-plane image resolution. The systemic error between CT and MRI (−0.29 mm[-0.26;-0.32]), indicated that the measurements were systematically smaller on the black bone MRI. Analysis after grouping by tooth type reduced the random error, while the systematic difference remained. For instance, greater differences were observed for the interdentium locations between the pre-molars and molars than for the incisors and canines (Figure 2).

We used formal method comparison statistics to compare black bone MRI and CT, as both imaging techniques are based on different physical phenomena (i.e. nuclear relaxivity for MRI, and photon absorption for CT). Both medical image acquisition methods inherently suffer from partial volume effects, which arise from the limited resolution of a scan; thus, when a tissue is only partly within a voxel, the intensity of that voxel is averaged between that and the adjacent tissue, resulting in blurring or smearing of the intensities. The in-plane resolution is better in MRI than in CT, resulting in sharper bone-tissue transition (Figure 1). Treece et al. showed that an intensity threshold method for cortical bone width measurements on CT overestimated the actual width by approximately 0.4 mm—as a result of the partial volume effect—if the cortical bone width was larger than approximately 1 mm [25]. The Hamada et al. method used in this study is a visual intensity threshold method. Therefore, a similar overestimation can be expected. This suggests that the smaller measurements on MRI may be closer to *in vivo* reality than those on CT, as our systematic error of –0.29 mm resembles the error of Treece et al. [25].

The sharper bone-tissue transition of the black bone MRI may also explain the significantly better inter-observer intra-class correlation (ICC) consistency for MRI, as the border of the bone may be more consistently placed. Nevertheless, MRI has its own challenges, which were also evident in this study. While CT scans can experience severe hyper-intense streaking distortions due to metal implants, they do not affect transverse slices without metal (i.e. inferior of the teeth level). Hence, the artifacts did not interfere with the cortical mandible bone intensities. In contrast, MRI suffers from magnetic field susceptibility artifacts, which refers to distortion of the magnetic field due to magnetization of a substance by the magnetic field itself [26]. This results in a three-dimensional elliptical hypo-dense signal on the MRI scan, which is particularly strong for some metals. This artifact inhibited some measurements for two patients (see an example in Appendix A3), while it did not in 4 other patients with metal artifacts on CT. Another concern is geometric distortion artifacts due to motion [27]. While motion can impact head and neck CT scans, which have sub-minute acquisition times, it is imperative to give concern in head and neck MRI scans due to longer scan times (∼5-8 minutes), creating extended opportunities for the patient to move during scan acquisition, especially if the patient is not fixated in a head and neck thermoplastic mask like in our study..

Our inter-observer analyses showed comparable variation in inter-observer measurements per Hamada et al. [12] between CT and MRI. The relatively poor inter-observer agreement and consistency for both imaging modalities demonstrates that the cortical bone measurement methodology is susceptible to observer variation, regardless of the image type. Iwata et al. also previously reported low inter-observer reliability between 2 observers measuring the cortical bone thickness with the Hamada approach in both MRONJ and non-MRONJ patients [3], and consequently could not recommend a reliable cut-off value for clinically meaningful cortical bone width change. Although the ICC was significantly higher for MRI in this study, the ICC was poor for both modalities in the 11 observers. The multi-observer measurements showed cluster type behavior (Figure 4), typifying a systematic bias among observers (Figure 4).

Qualitative evaluation of inter-observer analyses revealed that there are many factors that can influence the measurements of mandibular segmentation and morphometry. An example that leaves room for interpretation and flexibility are the oblique reconstruction of the scans to intersect the mandible perpendicularly, which requires scan angulated and rotated in the 3 directions. First, the sagittal and coronal planes need to be perpendicular to the mandible (blue and pink axes in Figure 2B,D) and which can lead to variation in interpretation as a function of slice selection alignment, which also alters localization of interdentium. A similar problem arises in the determining the transverse plane angulation (yellow axis in sagittal view Figure 2A,C) which needs to be parallel to the curving inferior mandible border and perpendicular position of the teeth roots. The final step is the rotation of the mandible cross-section (Figure 2E,F), which is often curved, leaving room for different interpretations of this rotation. These potential biases are cumulative in nature as a difference in the first step is propagated to all subsequent steps, altering the measurements. Moreover, measurements can also be different after cross-sectional positioning as the estimation of the mandible border will be determined differently based on the window/level of the intensity. While a standard window/level could be determined for CT, as proposed in this study (Appendix A2), MRI scans vary in intensity values with no standard window/level, which is clearly seen in highly variable maximum intensity values in Table 1. In this study, we have compared the intensity scaling on visual comparativeness. Standardization of MRI acquisition or window/level normalization is therefore a pre-requisite for consistent cortical bone measurements, as we performed in this series.

Future research should focus on automation of the Hamada et al. measurement in order to improve the standardization of the cortical bone measurements, as well as to improve clinical feasibility by reducing the time needed per measurement. Currently, a single patient single scan measurement takes about ∼ 2.5 hours, which is labor-intensive and thus leaves room for improvement. An alternative methodology may be to measure the width of the cortical bone over the entire surface of the mandible, which has shown potential for other anatomical sites [25], but this requires precise (auto-)contouring of the cortical bone. We believe that semi-automated or automated dental segmentation efforts may allow for more systematic morphometric assessment of dental structures [28] and may allow for more reproducible measures than human observers, and is thus an area ripe for future investigation.

## Conclusion

The black bone MR sequence shows promise as a high-resolution alternative to CT for cortical mandible bone measurements for monitoring and early diagnosis of osteonecrosis. Although MRI comes with its own challenges, the need for non-ionizing imaging techniques for cortical mandible bone thickness evaluation is crucial in order to allow for frequent monitoring of cortical bone changes. The inter-observer analyses showed significantly more consistent cortical bone measurements on black bone MRI than on CT; however, this was still relatively poor for both modalities, suggesting that the cortical bone measurement methodology is susceptible to inter-observer variation. In future studies, we intend to focus on reducing inter-observer susceptibility by standardization and preferably (semi-)automation of measuring the cortical mandible bone morphology. This may improve both the consistency as well as decrease the labor-intensive time needed to obtain the measurements.

## Data Availability

MRI and CT DICOM data & measurements will be available shortly on figshare: doi:10.6084/m9.figshare.12670754

https://doi.org/10.6084/m9.figshare.12670754

## References

[1] Imai Y, Hasegawa T, Takeda D, Kusumoto J, Akashi M, Ri S, et al. Evaluation and comparison of CT values in bisphosphonate-related osteonecrosis of the jaw. J Oral Maxillofac Surgery, Med Pathol 2016;28:19–25.

[2] Brown JP, Roux C, Ho PR, Bolognese MA, Hall J, Bone HG, et al. Denosumab significantly increases bone mineral density and reduces bone turnover compared with monthly oral ibandronate and risedronate in postmenopausal women who remained at higher risk for fracture despite previous suboptimal treatment with an oral bis. Osteoporos Int 2014;25:1953–61.

[3] Iwata E, Akashi M, Kishimoto M, Kusumoto J, Hasegawa T, Furudoi S, et al. Meaning and limitation of cortical bone width measurement with DentaScan in medication-related osteonecrosis of the jaws. Kobe J Med Sci 2016;62:E114–9.

[4] Sciubba JJ, Goldenberg D. Oral complications of radiotherapy. Lancet Oncol 2006;7:175–83.

[5] Kuo TJ, Leung CM, Chang HS, Wu CN, Chen WL, Chen GJ, et al. Jaw osteoradionecrosis and dental extraction after head and neck radiotherapy: A nationwide population-based retrospective study in Taiwan. Oral Oncol 2016;56:71–7.

[6] Hamilton JD, Lai SY, Ginsberg LE. Superimposed infection in mandibular osteoradionecrosis: Diagnosis and outcomes. J Comput Assist Tomogr 2012;36:725–31.

[7] Mohamed ASR, Hobbs BP, Hutcheson KA, Murri MS, Garg N, Song J, et al. Dose-volume correlates of mandibular osteoradionecrosis in Oropharynx cancer patients receiving intensity-modulated radiotherapy: Results from a case-matched comparison. Radiother Oncol 2017;124:232–9.

[8] DeNardo DG, Brennan DJ, Rexhepaj E, Ruffell B, Shiao SL, Madden SF, et al. Leukocyte Complexity Predicts Breast Cancer Survival and Functionally Regulates Response to Chemotherapy. Cancer Discov 2011;1:54–67.

[9] Khan AA, Morrison A, Hanley DA, Felsenberg D, McCauley LK, O’Ryan F, et al. Diagnosis and management of osteonecrosis of the jaw: A systematic review and international consensus. J Bone Miner Res 2015;30:3–23.

[10] Chronopoulos A, Zarra T, Ehrenfeld M, Otto S. Osteoradionecrosis of the jaws: definition, epidemiology, staging and clinical and radiological findings. A concise review. Int Dent J 2018;68:22–30.

[11] Smith-Bindman R. Radiation Dose Associated With Common Computed Tomography Examinations and the Associated Lifetime Attributable Risk of Cancer. Arch Intern Med 2009;169:2078.

[12] Hamada H, Matsuo A, Koizumi T, Satomi T, Chikazu D. A simple evaluation method for early detection of bisphosphonate-related osteonecrosis of the mandible using computed tomography. J Cranio-Maxillofacial Surg 2014;42:924–9.

[13] Eley KA, Watt-Smith SR, Golding SJ. “Black bone” MRI: A potential alternative to CT when imaging the head and neck: Report of eight clinical cases and review of the Oxford experience. Br J Radiol 2012;85:1457–64.

[14] Eley KA, Mcintyre AG, Watt-Smith SR, Golding SJ. “Black bone” MRI: A partial flip angle technique for radiation reduction in craniofacial imaging. Br J Radiol 2012;85:272–8.

[15] Eley KA, Watt-Smith SR, Sheerin F, Golding SJ. “Black Bone” MRI: a potential alternative to CT with three-dimensional reconstruction of the craniofacial skeleton in the diagnosis of craniosynostosis. Eur Radiol 2014;24:2417–26.

[16] Dremmen MHG, Wagner MW, Bosemani T, Tekes A, Agostino D, Day E, et al. Does the addition of a “black bone” sequence to a fast multisequence trauma mr protocol allow MRI to replace ct after traumatic brain injury in children? Am J Neuroradiol 2017;38:2187–92.

[17] Eley KA, Watt-Smith SR, Golding SJ. “black Bone” MRI: A novel imaging technique for 3D printing. Dentomaxillofacial Radiol 2017;46.

[18] Kralik SF, Supakul N, Wu IC, Delso G, Radhakrishnan R, Ho CY, et al. Black bone MRI with 3D reconstruction for the detection of skull fractures in children with suspected abusive head trauma. Neuroradiology 2019;61:81–7.

[19] Martin Bland J, Altman DG. Statistical methods for assessing agreement between two methods of clinical measurement. Lancet 1986;38:97–107.

[20] Lin LI. A Concordance Correlation Coefficient to Evaluate Reproducibility Author (s): Lawrence I-Kuei Lin Published by?: International Biometric Society Stable URL?: http://www.jstor.org/stable/2532051 REFERENCES Linked references are available on JSTOR for thi. Biomatrics 1989;45:255–68.

[21] Payne RB. Method comparison: Evaluation of least squares, Deming and Passing/Bablok regression procedures using computer simulation. Ann Clin Biochem 1997;34:319–20.

[22] Linnet K. Performance of Deming regression analysis in case of misspecified analytical error ratio in method comparison studies. Clin Chem 1998;44:1024–31.

[23] Koo TK, Li MY. A Guideline of Selecting and Reporting Intraclass Correlation Coefficients for Reliability Research. J Chiropr Med 2016;15:155–63.

[24] Shrout PE, Fleiss JL. Intraclass correlations: Uses in assessing rater reliability. Psychol Bull 1979;86:420–8.

[25] Treece G, Gee A. Cortical Bone Mapping: Measurement and Statistical Analysis of Localised Skeletal Changes. Curr Osteoporos Rep 2018;16:617–25.

[26] Schenck JF. The role of magnetic susceptibility in magnetic resonance imaging: MRI magnetic compatibility of the first and second kinds. Med Phys 1996;23:815–50.

[27] Zaitsev M, Maclaren J, Herbst M. Motion artifacts in MRI: A complex problem with many partial solutions. J Magn Reson Imaging 2015;42:887–901.

[28] Thariat J, Ramus L, Maingon P, Odin G, Gregoire V, Darcourt V, et al. Dentalmaps: Automatic dental delineation for radiotherapy planning in head-and-neck cancer. Int J Radiat Oncol Biol Phys 2012;82:1858–65.

